# ICU Hour-24 Landmark Prediction of In-Hospital Mortality in Critically Ill Patients With Coronary Artery Disease: Development in MIMIC-IV and External Validation in eICU

**DOI:** 10.64898/2026.07.28.26359081

**Authors:** Sakshie Pathak, Janet Sanjaya, Yong Si, Mohammadsaeed Haghi, Nausin Kudrot, Greg Placencia, Kamiar Alaei, Maryam Pishgar

## Abstract

**Background:** Retrospective intensive care unit prediction models are vulnerable to temporal leakage when predictors include information recorded after the intended prediction time. We developed and externally validated an ICU hour-24 landmark prediction pipeline for in-hospital mortality among critically ill patients with coronary artery disease using timestamp-restricted features from MIMIC-IV and eICU.

**Methods:** Adults with coronary artery disease or coronary heart disease who were alive and remained under ICU observation at hour 24 were included. MIMIC-IV was used for model development, and eICU was reserved for external validation. Dynamic events were restricted to ICU admission through hour 24 in MIMIC-IV and offsets of 0–1440 minutes in eICU before aggregation. We evaluated an XGBoost model using baseline and respiratory-support predictors and a 102-predictor random forest using baseline, respiratory-support, and treatment predictors. Robustness was assessed across 30 repeated patient-grouped MIMIC-IV validation splits, and external uncertainty was estimated using 1,000 subject-clustered bootstrap resamples.

**Results:** The MIMIC-IV cohort included 4,341 ICU stays with 993 deaths, and the eICU cohort included 19,464 stays with 2,237 deaths. In eICU, XGBoost achieved a ROC-AUC of 0.7973 (95% CI, 0.7878–0.8068), a PR-AUC (calculated as average precision) of 0.3627 (95% CI, 0.3423–0.3843), and a Brier score of 0.1189 (95% CI, 0.1164–0.1213). The random forest achieved a ROC-AUC of 0.8060 (95% CI, 0.7960–0.8154), a PR-AUC of 0.3687 (95% CI, 0.3471–0.3915), and a Brier score of 0.1032 (95% CI, 0.1012–0.1054). The random forest had a modestly higher ROC-AUC and lower Brier score than XGBoost. Mean validation ROC-AUCs across repeated MIMIC-IV splits were 0.7970 and 0.7954, respectively. Exploratory analyses suggested that narrower, consistently harmonized feature sets transported more reliably than broader expansions.

**Conclusions:** Timestamp-restricted first-day models achieved external ROC-AUCs of approximately 0.80. However, external calibration remained imperfect, and local recalibration and prospective evaluation would be required before clinical use.

## 1 Background

Coronary artery disease (CAD) and coronary heart disease (CHD) remain major contributors to cardiovascular morbidity and mortality worldwide [1, 2]. Patients with coronary disease who require intensive care represent a high-risk population and may have concurrent shock, respiratory failure, renal dysfunction, sepsis, metabolic derangement, and complex treatment exposure. During the first 24 hours of intensive care unit (ICU) admission, clinicians make decisions regarding monitoring intensity, ventilatory support, vasoactive therapy, transfer, and escalation of care. Prediction models intended to support risk stratification at this stage should therefore use only information available by the specified prediction time.

Large critical-care databases have enabled the development and evaluation of machine-learning models for mortality and other adverse clinical outcomes [3–6]. Machine-learning approaches have also been applied to cardiovascular risk prediction, including stroke prediction in patients with revascularized coronary artery disease and early mortality risk assessment in critically ill patients with coexisting cardiovascular conditions [7, 8]. However, retrospective ICU model development requires careful alignment of the prediction time, predictor window, and outcome period. Predictors extracted from complete ICU or hospital stays may include information recorded after the intended prediction landmark, while administrative and diagnosis variables may reflect information documented later during hospitalization or after discharge. Dynamic measurements may also differ across databases in their timing, frequency, and documentation practices. These factors can introduce temporal leakage and produce performance estimates that do not accurately represent the intended clinical prediction task [9–11].

Landmark analysis addresses part of this problem by defining a specific prediction time and restricting the prediction population to patients who remain alive and under observation at that time [12, 13]. In the present study, the landmark was ICU hour 24. The resulting prediction task estimates subsequent in-hospital mortality among patients who survived and remained under ICU observation through the first 24 hours. It should therefore not be interpreted as an admission-time mortality prediction or triage model.

Recent disease-specific ICU mortality studies have examined dynamic or temporal feature construction in patients with myocardial infarction, alcoholic cirrhosis, and abdominal aortic aneurysm [14–16]. However, the cross-database behavior of temporally restricted feature families remains less clearly characterized. In the present study, respiratory-support and treatment predictors were reconstructed from raw event tables using a fixed hour-24 information boundary. Models were developed in MIMIC-IV and subsequently evaluated in eICU, with multiple feature families examined to characterize their performance across the two databases.

Accordingly, this study aimed to develop and externally evaluate an ICU hour-24 mortality prediction pipeline for critically ill patients with coronary disease using timestamp-restricted first-day information. We specifically examined whether respiratory-support and treatment predictors retained predictive value when transported from MIMIC-IV to eICU and whether broader feature expansion was associated with consistent cross-database performance. In addition to discrimination, we evaluated calibration, probabilistic accuracy, and model stability to characterize the limitations of transferring predicted risks between the two databases.

## 2 Methods

### 2.1 Study design and data sources

We conducted a retrospective model-development and external-validation study using de-identified intensive care data from MIMIC-IV and the eICU Collaborative Research Database [17–19]. The prediction landmark was 24 hours after ICU admission. MIMIC-IV was used for cohort construction, model development, internal validation, repeated patient-grouped robustness analysis, and timestamp-ordered testing. eICU was used as the external validation cohort.

eICU data were not used for model fitting, hyperparameter tuning, or initial feature-family selection. After the final model specifications had been defined using MIMIC-IV, the models were refit on the full MIMIC-IV analysis cohort and applied unchanged to eICU. Additional feature-family comparisons in eICU were conducted after the primary external benchmark and were treated as exploratory transport analyses rather than as a basis for model selection.

### 2.2 Study population and cohort construction

The unit of analysis was an eligible adult ICU stay. We included ICU stays of patients aged 18 years or older with coronary artery disease or coronary heart disease identified using the diagnosis-code criteria applied in each database. Eligible stays were required to have an ICU length of stay of at least 24 hours, valid timing information required to define the hour-24 landmark, and an available in-hospital mortality outcome.

Because prediction was performed at ICU hour 24, patients were required to be alive and remain under ICU observation at the landmark. Stays ending before hour 24, including those in which death occurred before the landmark, were therefore excluded from the analysis cohort.

### 2.3 Outcome and prediction landmark

The outcome was in-hospital mortality occurring after the ICU hour-24 landmark. Predictor information was restricted to data available from ICU admission through hour 24, and the models estimated subsequent in-hospital mortality risk after the landmark among patients who were alive and remained in the ICU at the prediction time.

This design separated deaths occurring before the landmark from the subsequent mortality prediction task. Accordingly, the models do not apply to patients who died or left the ICU before hour 24 and should not be interpreted as admission-time mortality prediction or triage models.

### 2.4 Predictor construction and temporal eligibility

Candidate predictors were organized into five domains: demographics and baseline descriptors; bedside and laboratory measurements; derived physiologic variables and interaction terms; respiratory-support summaries; and treatment indicators and event counts.

The baseline feature set included bedside, laboratory, and derived predictors available at or before the ICU hour-24 landmark. Respiratory-support features summarized ventilatory and respiratory events, including positive end-expiratory pressure, respiratory-rate settings or observations, tidal-volume variables, mean airway pressure, and event counts. Where applicable, dynamic measurements were summarized using the first, last, minimum, maximum, mean, count, and change observed during the feature window. Treatment exposure was represented using binary indicators and counts of harmonized treatment events recorded by hour 24.

All dynamic events were restricted to the prespecified feature window before aggregation. For MIMIC-IV, events were retained only when

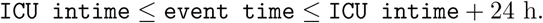

For eICU, events were retained only when

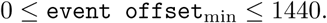

Database-specific event names were mapped to harmonized feature groups, and summary features were constructed only after application of the corresponding timestamp filters. The aggregated feature tables were then left-joined to the eligible analysis cohorts so that all qualifying ICU stays were retained. Absence of a recorded support or treatment event was encoded as zero for binary indicators and event counts. Missing numeric summaries, such as unavailable ventilator settings, were retained as missing values and addressed during preprocessing.

### 2.5 Missing data and preprocessing

For each MIMIC-IV development split, numeric predictors were imputed using medians estimated from the corresponding training data. Categorical predictors, when present, were imputed using modes estimated from the same fitting data. Missingness-indicator variables were generated for numeric predictors during preprocessing.

For final external validation, preprocessing parameters were re-estimated from the full MIMIC-IV analysis cohort. The resulting MIMIC-IV-derived imputation parameters and fitted preprocessing pipeline were then applied unchanged to eICU. Tree-based models were fitted without feature scaling.

### 2.6 Feature-family specifications

Six feature-family specifications were examined across the MIMIC-IV development analyses and the subsequent exploratory transport analyses. These included baseline predictors alone; baseline predictors with respiratory-support features; baseline predictors with medication or treatment exposure; baseline predictors with vital-sign extensions; baseline predictors with the full set of support and treatment extensions; and a focused baseline, respiratory-support, and treatment specification. The number of predictors in each feature family is shown in Table 1.

**Table 1.** Feature-family specifications and number of predictors.

| Feature family | Number of predictors |
| --- | --- |
| Baseline predictors | 38 |
| Baseline + respiratory support | 96 |
| Baseline + medication/treatment exposure | 57 |
| Baseline + vital-sign extensions | 65 |
| Baseline + all support/treatment features | 142 |
| Baseline + respiratory support + treatment | 102 |

### 2.7 Model development and final fitting

Candidate algorithms included extreme gradient boosting (XGBoost) and random forests [20, 21]. Model development and specification decisions were based on MIMIC-IV analyses. The XGBoost model using baseline and respiratory-support predictors was selected according to the MIMIC-IV validation ROC-AUC criterion. A 102-predictor random-forest specification using baseline, respiratory-support, and treatment predictors was retained as a secondary MIMIC-IV-defined model because it showed comparable performance during repeated patient-grouped validation. eICU performance was not used for model fitting or hyperparameter selection.

For external evaluation, both model specifications were refitted using all eligible MIMIC-IV analysis stays. Imputation parameters were re-estimated from the full MIMIC-IV refit cohort, after which the fitted preprocessing pipelines and models were applied unchanged to the eICU cohort. The final model specifications and training settings are summarized in Table 2.

**Table 2.**
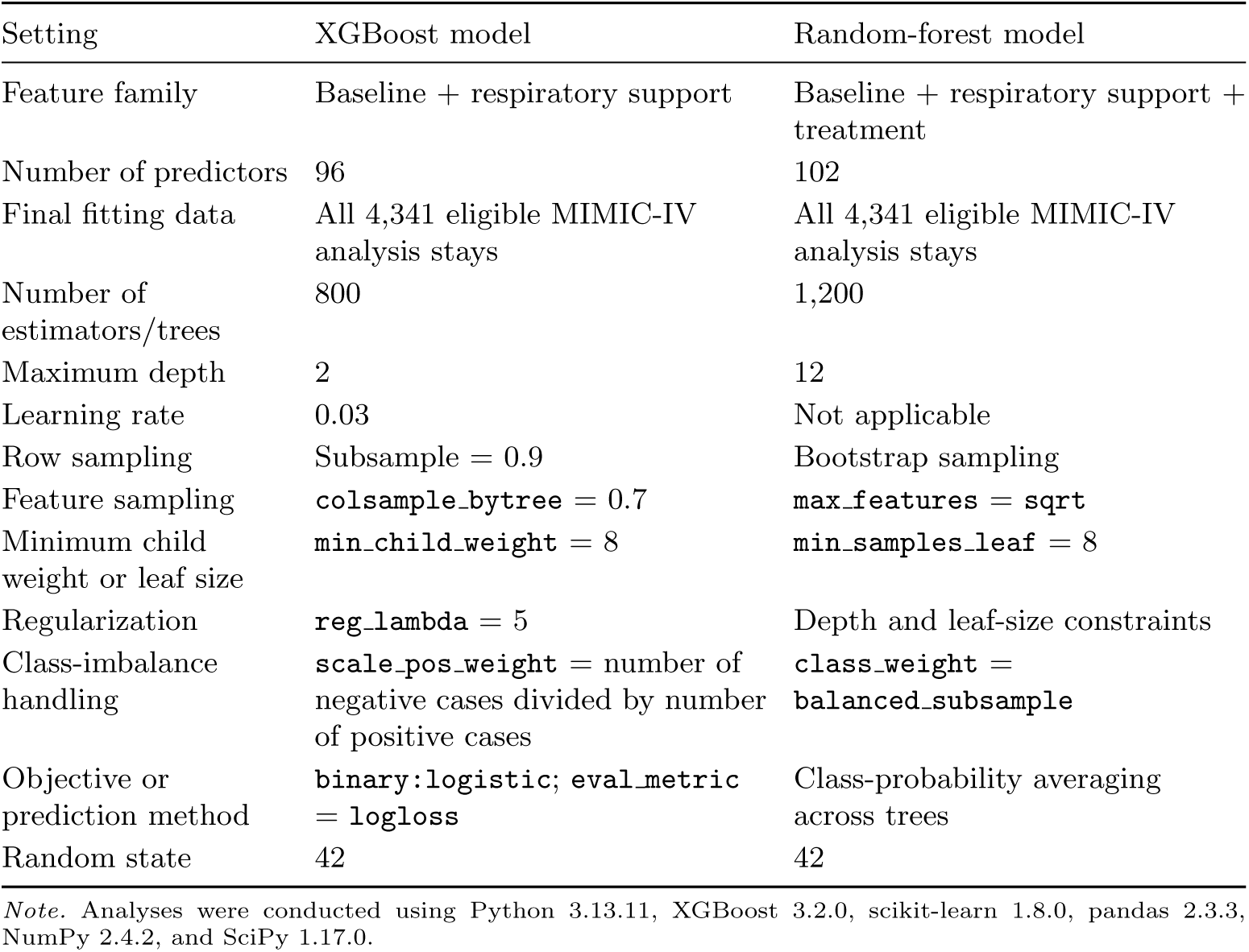
Final model specifications and training settings.

| Setting | XGBoost model | Random-forest model |
| --- | --- | --- |
| Feature family | Baseline + respiratory support | Baseline + respiratory support + treatment |
| Number of predictors | 96 | 102 |
| Final fitting data | All 4,341 eligible MIMIC-IV analysis stays | All 4,341 eligible MIMIC-IV analysis stays |
| Number of estimators/trees | 800 | 1,200 |
| Maximum depth | 2 | 12 |
| Learning rate | 0.03 | Not applicable |
| Row sampling | Subsample = 0.9 | Bootstrap sampling |
| Feature sampling | `colsample_bytree = 0.7` | `max_features = sqrt` |
| Minimum child weight or leaf size | `min_child_weight = 8` | `min_samples_leaf = 8` |
| Regularization | `reg_lambda = 5` | Depth and leaf-size constraints |
| Class-imbalance handling | `scale_pos_weight =` number of negative cases divided by number of positive cases | `class_weight = balanced_subsample` |
| Objective or prediction method | `binary:logistic`; `eval_metric = logloss` | Class-probability averaging across trees |
| Random state | 42 | 42 |

*Note.* Analyses were conducted using Python 3.13.11, XGBoost 3.2.0, scikit-learn 1.8.0, pandas 2.3.3, NumPy 2.4.2, and SciPy 1.17.0.

### 2.8 Internal validation and robustness assessment

MIMIC-IV data were partitioned at the subject level so that all eligible ICU stays belonging to the same patient remained within the same subset. For the primary development partition, 20% of subjects were first reserved for the held-out internal test set. The remaining 80% of subjects were divided into training and validation subsets using a 75:25 allocation. This produced an approximate overall allocation of 60% training, 20% validation, and 20% internal testing. Model and feature-family decisions were based on the MIMIC-IV development analyses.

A separate subject-level timestamp-ordered MIMIC-IV split was created using each patient’s first eligible ICU admission time, defined as the minimum ICU intime across that patient’s eligible stays. Subjects were ordered by this first admission time and then by subject id. The earliest 70% of subjects were assigned to the temporal training set and the latest 30% to the temporal test set. All eligible ICU stays belonging to each subject were retained within the corresponding partition.

To assess dependence on a single favorable patient-grouped partition, the same patient-grouped splitting procedure was repeated 30 times. Within each repeat, pre- processing and model fitting were performed using the corresponding training subset, and performance was evaluated on the corresponding validation subset. Training ROC-AUC, validation ROC-AUC, validation PR-AUC, and validation Brier score were summarized across the 30 repeats.

### 2.9 Performance measures and statistical analysis

Reporting was structured with reference to established guidance for clinical prediction-model studies [22–24]. Model performance was assessed using measures of discrimination, probabilistic accuracy, calibration, and potential clinical utility [25–27].

Discrimination was evaluated using the area under the receiver operating characteristic curve (ROC-AUC) and precision–recall performance. The reported PR-AUC values corresponded to average precision, calculated using `sklearn.metrics.average precision score`. Precision–recall curves were generated separately using `sklearn.metrics.precision recall curve`. PR-AUC was included because mortality represented a minority of observations in the external evaluation cohort [28, 29]. Probabilistic accuracy was evaluated using the Brier score, with lower values indicating lower mean squared prediction error [30].

External uncertainty was estimated using 1,000 subject-clustered bootstrap resamples of the eICU cohort. Subjects were sampled with replacement, and all ICU stays associated with each sampled subject were retained within the corresponding bootstrap replicate. Ninety-five percent confidence intervals were obtained from the empirical 2.5th and 97.5th percentiles of the bootstrap distributions.

Because the two final models were evaluated on the same eICU cohort, differences in ROC-AUC and Brier score were calculated within paired subject-clustered bootstrap replicates [31]. The paired bootstrap sign-test p-value for the ROC-AUC comparison was calculated using a plus-one correction. The Brier-score comparison was summarized using the paired bootstrap confidence interval.

Calibration was assessed using calibration intercept and calibration slope estimated from a logistic calibration model,

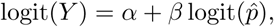

where predicted probabilities were clipped before logit transformation to avoid infinite values. Calibration curves were constructed using equal-frequency predicted-risk groups [32].

Decision-curve analysis was used to estimate net benefit across threshold probabilities from 0.01 to 0.50 [33]. Model net benefit was compared with treat-all and treat-none strategies. Because no clinical intervention or decision threshold was prespecified, decision-curve results were treated as exploratory.

Feature importance was calculated separately for the two tree-based models using the native `feature importances` values returned by the fitted estimators. Importance values for missingness-indicator columns generated during preprocessing were aggregated with their corresponding parent predictors. These analyses were interpreted as model-specific importance rankings rather than as signed associations, causal effects, or SHAP explanations.

### 2.10 Exploratory subgroup and distribution-shift analyses

External subgroup discrimination was examined for the XGBoost respiratory-support model across four sets of categories: age younger than 65 years versus 65 years or older, male versus female sex, presence versus absence of a recorded respiratory-support event, and presence versus absence of a treatment flag. ROC-AUC was calculated within each subgroup, with bootstrap confidence intervals used to summarize uncertainty. These analyses were considered exploratory and were not used for model selection.

Cross-database feature differences were explored for selected predictors using the population stability index (PSI), the absolute standardized mean difference, and the absolute difference in missingness proportion between MIMIC-IV and eICU. These measures were used descriptively to identify variables with differences in distribution or documentation across the two databases; no formal drift threshold was used for model exclusion or selection.

Predicted-risk distributions were also compared descriptively across the MIMIC-IV validation cohort, the timestamp-ordered MIMIC-IV test cohort, and the eICU evaluation cohort. The subgroup, feature-drift, and predicted-risk distribution analyses were performed after the primary model-evaluation workflow and were interpreted as exploratory assessments of model behavior and cross-database transport.

Detailed pseudocode for timestamp-restricted hour-24 feature construction and the model-development and evaluation workflow is provided in Supplementary Algorithms S1 and S2.

## 3 Results

### 3.1 Study population

The MIMIC-IV development cohort comprised 4,341 ICU stays from 3,982 unique patients, including 993 in-hospital deaths, corresponding to a mortality prevalence of 22.9%. The patient-grouped development partition included 2,603 training stays, 870 validation stays, and 868 held-out internal test stays. The separate timestamp-ordered MIMIC-IV analysis included 3,053 training stays and 1,288 test stays.

The eICU external evaluation cohort comprised 19,464 ICU stays, including 2,237 in-hospital deaths, corresponding to a mortality prevalence of 11.49%. Cohort eligibility criteria and final cohort sizes are summarized in Figure 2.

**Fig. 1.**
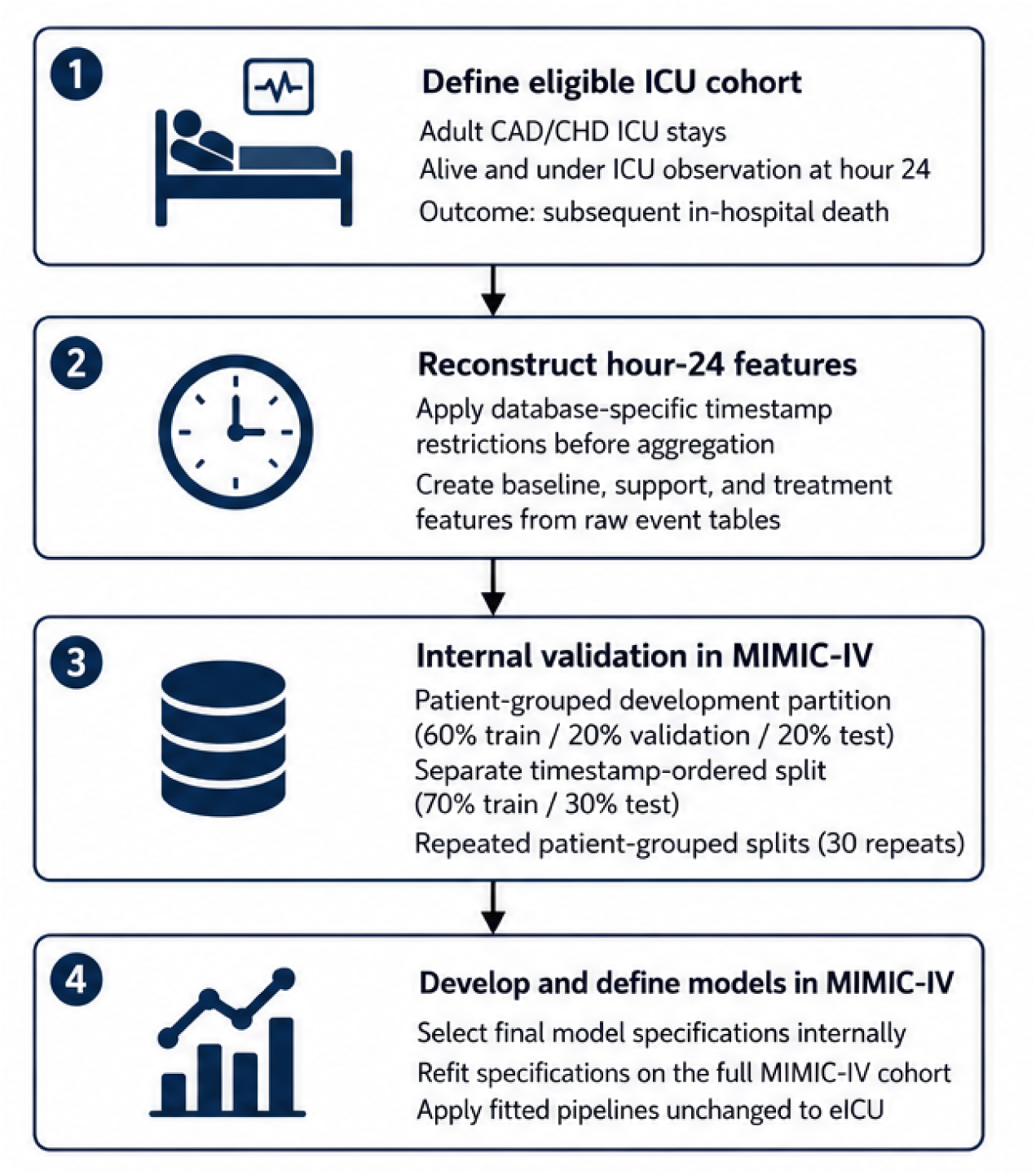
Overview of the ICU hour-24 prediction workflow. Eligible patients were required to be alive and under ICU observation at the hour-24 landmark. Database-specific timestamp restrictions were applied before aggregation of dynamic predictors. Model development, patient-grouped validation, and timestamp-ordered testing were performed in MIMIC-IV. The final preprocessing pipelines and model specifications were subsequently refitted on the full MIMIC-IV cohort and applied unchanged to eICU.

**Fig. 2.**
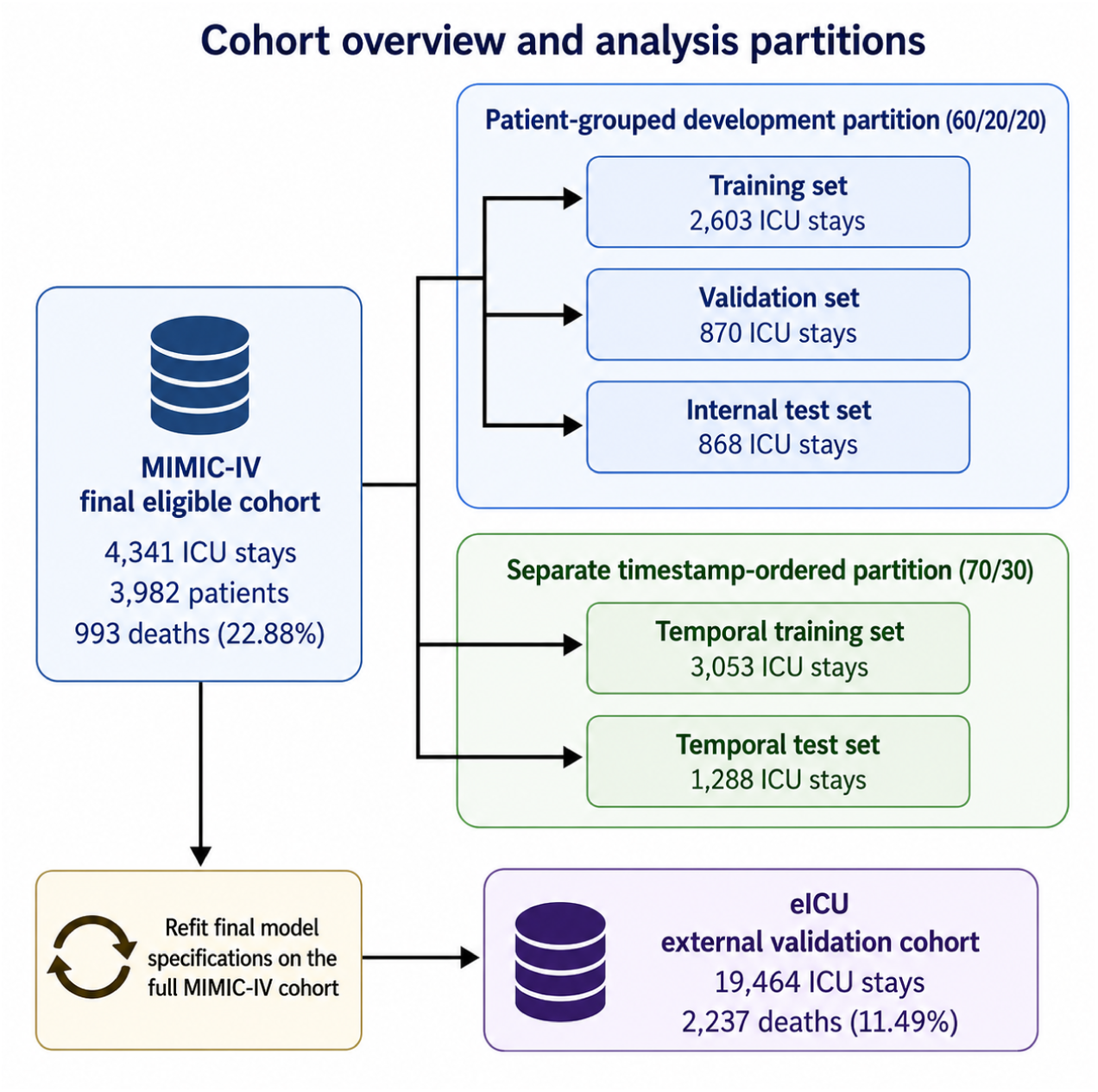
Cohort overview and analysis partitions. Both databases were restricted to eligible adult CAD/CHD ICU stays in which patients were alive and remained under ICU observation at hour 24. The MIMIC-IV cohort was evaluated using a patient-grouped development partition and a separate subject-level timestamp-ordered partition. Final model specifications were refitted on the full MIMIC-IV cohort and externally validated in eICU.

### 3.2 External performance in eICU

External performance estimates for the two final model specifications are presented in Table 3. The XGBoost model using baseline and respiratory-support predictors achieved a ROC-AUC of 0.7973 (95% CI, 0.7878–0.8068), a PR-AUC of 0.3627 (95% CI, 0.3423–0.3843), and a Brier score of 0.1189 (95% CI, 0.1164–0.1213).

**Table 3.** External performance in the eICU evaluation cohort with subject-clustered bootstrap 95% confidence intervals.

**Panel A. Discrimination and probabilistic accuracy**
| Model | Feature specification | ROC-AUC<br>(95% CI) | PR-AUC (95%<br>CI) | Brier score<br>(95% CI) |
| --- | --- | --- | --- | --- |
| XGBoost | Baseline + respiratory support | 0.7973<br>(0.7878–0.8068) | 0.3627<br>(0.3423–0.3843) | 0.1189<br>(0.1164–0.1213) |
| Random forest | Baseline + respiratory support + treatment | 0.8060<br>(0.7960–0.8154) | 0.3687<br>(0.3471–0.3915) | 0.1032<br>(0.1012–0.1054) |

**Panel B. Calibration**
| Model | Feature specification | Calibration intercept<br>(95% CI) | Calibration slope<br>(95% CI) |
| --- | --- | --- | --- |
| XGBoost | Baseline + respiratory support | −1.2732<br>(−1.3264 to −1.2190) | 1.0318<br>(0.9863–1.0779) |
| Random forest | Baseline + respiratory support + treatment | −0.3561<br>(−0.4370 to −0.2734) | 1.7750<br>(1.6928–1.8586) |

*Note.* CI, confidence interval; PR-AUC, precision–recall performance summarized using average precision; ROC-AUC, area under the receiver operating characteristic curve. Confidence intervals were estimated using 1,000 subject-clustered bootstrap resamples of the eICU cohort.

The random-forest model using baseline, respiratory-support, and treatment predictors achieved a ROC-AUC of 0.8060 (95% CI, 0.7960–0.8154), a PR-AUC of 0.3687 (95% CI, 0.3471–0.3915), and a Brier score of 0.1032 (95% CI, 0.1012–0.1054). The no-skill precision reference was 0.1149, corresponding to the mortality prevalence in the eICU cohort.

In paired subject-clustered bootstrap comparisons, the random forest had a modestly higher ROC-AUC than XGBoost, with a difference of 0.0087 (95% CI, 0.0036–0.0136; paired bootstrap sign-test *p <* 0.002). The random forest also had a lower Brier score, with a paired difference of *−*0.0156 (95% CI, *−*0.0171 to *−*0.0142; Table 4). External ROC and precision–recall curves are shown in Figure 3.

**Fig. 3.**
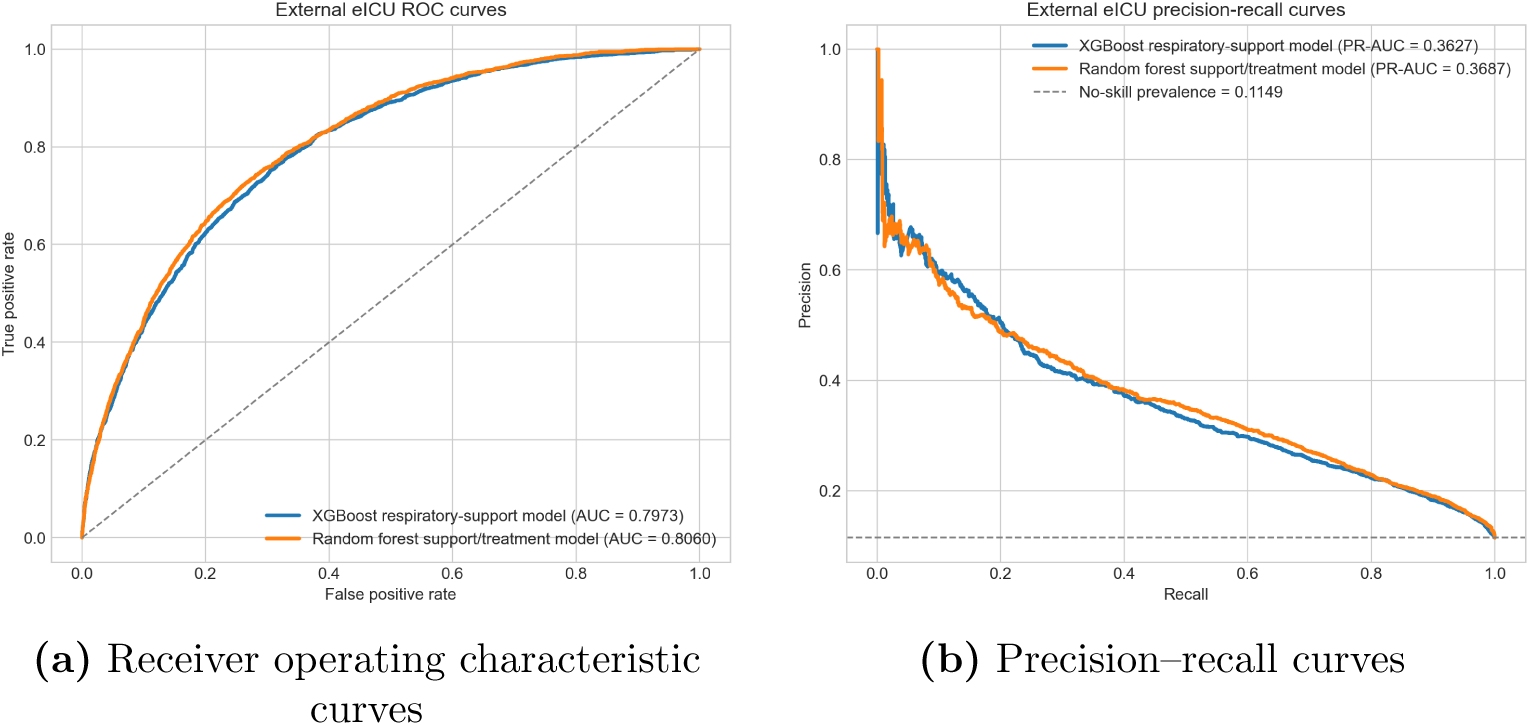
External discrimination in the eICU evaluation cohort. Receiver operating characteristic curves are shown in panel **(a)**, and precision–recall curves are shown in panel **(b)**. The no-skill reference in the precision–recall panel corresponds to the eICU mortality prevalence of 11.49%.

**Table 4.** Paired subject-clustered bootstrap comparison of the random-forest and XGBoost models in eICU.

| Metric | Random forest minus XGBoost | 95% CI |
| --- | --- | --- |
| ROC-AUC | 0.0087 | 0.0036 to 0.0136 |
| Brier score | −0.0156 | −0.0171 to −0.0142 |

*Note.* The paired bootstrap sign-test yielded $p < 0.002$ for the ROC-AUC comparison. Negative Brier-score differences favor the random-forest model.

### 3.3 Repeated patient-grouped robustness assessment

Performance across the 30 repeated patient-grouped MIMIC-IV validation splits is summarized in Table 5. The XGBoost model achieved a mean validation ROC-AUC of 0.7970 (SD, 0.0126), with an empirical interval of 0.7769–0.8171. The random-forest model achieved a mean validation ROC-AUC of 0.7954 (SD, 0.0120), with an empirical interval of 0.7733–0.8157.

**Table 5.** Performance across 30 repeated patient-grouped MIMIC-IV validation splits.

A. ROC-AUC performance
| Model | Feature specification | Mean<br>training<br>ROC-AUC | Mean<br>validation<br>ROC-AUC | SD | Empirical<br>interval |
| --- | --- | --- | --- | --- | --- |
| XGBoost | Baseline +<br>respiratory support | 0.9028 | 0.7970 | 0.0126 | 0.7769–0.8171 |
| Random<br>forest | Baseline +<br>respiratory support<br>+ treatment | 0.9786 | 0.7954 | 0.0120 | 0.7733–0.8157 |

**Panel B. Validation PR-AUC and Brier score**
| Model | Feature specification | Mean validation<br>PR-AUC | Mean validation<br>Brier score |
| --- | --- | --- | --- |
| XGBoost | Baseline + respiratory support | 0.5386 | 0.1722 |
| Random<br>forest | Baseline + respiratory support<br>+ treatment | 0.5318 | 0.1581 |

*Note.* Values are summaries across 30 repeated patient-grouped MIMIC-IV training and validation splits. The empirical interval summarizes the observed validation ROC-AUC distribution across repeated splits and should not be interpreted as a formal confidence interval. PR-AUC, precision–recall performance summarized using average precision; ROC-AUC, area under the receiver operating characteristic curve; SD, standard deviation.

The mean validation ROC-AUC difference between the two specifications was 0.0016, which was small relative to the variation observed across splits. The XGBoost and random-forest models achieved mean validation PR-AUCs of 0.5386 and 0.5318, respectively. Mean validation Brier scores were 0.1722 for XGBoost and 0.1581 for the random forest.

Apparent training discrimination exceeded validation discrimination for both models. Mean training ROC-AUCs were 0.9028 for XGBoost and 0.9786 for the random forest, corresponding to approximate training–validation gaps of 0.106 and 0.183, respectively.

### 3.4 Exploratory feature-family transport analyses

Exploratory feature-family results are presented in Tables 6 and 7. These analyses were conducted after the primary external benchmark and were used to characterize the behavior of related predictor families across MIMIC-IV and eICU.

**Table 6.**
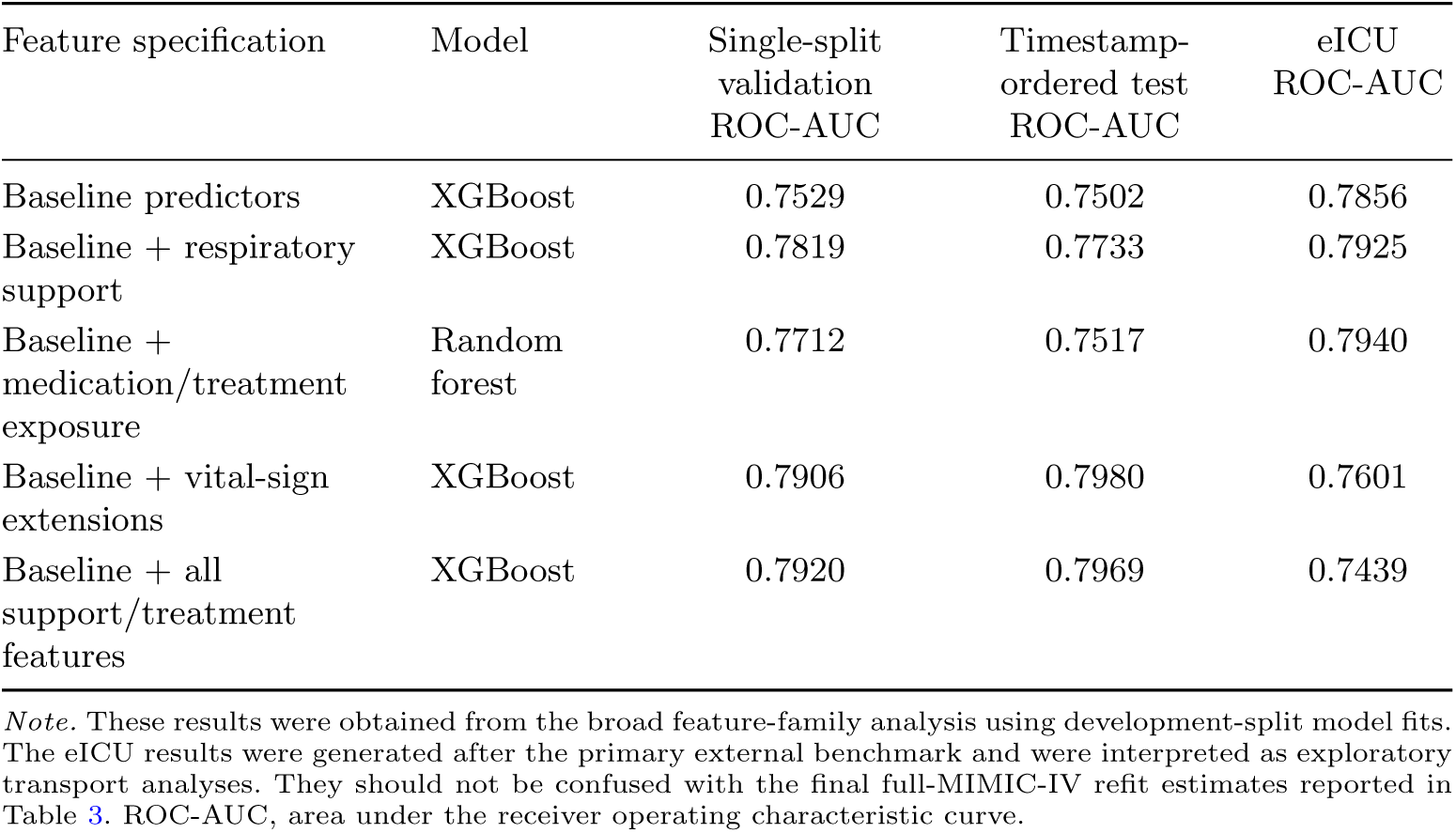
Exploratory cross-database performance of broad feature-family specifications.

| Feature specification | Model | Single-split<br>validation<br>ROC-AUC | Timestamp-ordered test<br>ROC-AUC | eICU<br>ROC-AUC |
| --- | --- | --- | --- | --- |
| Baseline predictors | XGBoost | 0.7529 | 0.7502 | 0.7856 |
| Baseline + respiratory support | XGBoost | 0.7819 | 0.7733 | 0.7925 |
| Baseline + medication/treatment exposure | Random forest | 0.7712 | 0.7517 | 0.7940 |
| Baseline + vital-sign extensions | XGBoost | 0.7906 | 0.7980 | 0.7601 |
| Baseline + all support/treatment features | XGBoost | 0.7920 | 0.7969 | 0.7439 |

*Note.* These results were obtained from the broad feature-family analysis using development-split model fits. The eICU results were generated after the primary external benchmark and were interpreted as exploratory transport analyses. They should not be confused with the final full-MIMIC-IV refit estimates reported in Table 3. ROC-AUC, area under the receiver operating characteristic curve.

**Table 7.** Exploratory cross-database performance of focused support, treatment, and medication feature specifications.

| Feature specification | Model | Single-split<br>validation<br>ROC-AUC | Timestamp-ordered test<br>ROC-AUC | eICU<br>ROC-AUC |
| --- | --- | --- | --- | --- |
| Baseline + respiratory support + treatment | Random forest | 0.7782 | 0.7730 | 0.8060 |
| Respiratory support + medication + treatment | Random forest | 0.7816 | 0.7672 | 0.8052 |
| Respiratory support + medication | Random forest | 0.7779 | 0.7660 | 0.8051 |
| Respiratory support + medication + RR/temperature | Random forest | 0.7802 | 0.7703 | 0.8045 |

*Note.* These focused feature-family analyses were conducted after the primary external benchmark and were used to characterize the cross-database behavior of related predictor combinations. The baseline, respiratory-support, and treatment row produced the same eICU ROC-AUC, to four decimal places, as the final random-forest model; however, the values in this table originate from the exploratory feature-family workflow, whereas Table 3 reports the final full-MIMIC-IV refit evaluation. RR, respiratory rate; ROC-AUC, area under the receiver operating characteristic curve.

In the broad feature-family comparison, the baseline XGBoost specification achieved an eICU ROC-AUC of 0.7856. Adding respiratory-support summaries increased the eICU ROC-AUC to 0.7925, while the baseline medication/treatment specification achieved an eICU ROC-AUC of 0.7940. In contrast, specifications incorporating broader vital-sign extensions or the full set of support and treatment extensions achieved eICU ROC-AUCs of 0.7601 and 0.7439, respectively, despite stronger performance in the MIMIC-IV validation or timestamp-ordered test analyses. Among the focused random-forest specifications, the baseline, respiratory-support, and treatment feature set achieved the highest eICU ROC-AUC of 0.8060. Adding medication variables produced similar but slightly lower eICU ROC-AUCs, ranging from 0.8045 to 0.8052. Overall, the exploratory comparisons showed that broader feature expansion did not consistently improve performance across the two databases.

### 3.5 External calibration

External calibration estimates are reported in Table 3 and illustrated in Figure 4. The XGBoost model had a calibration intercept of *−*1.2732 (95% CI, *−*1.3264 to *−*1.2190) and a calibration slope of 1.0318 (95% CI, 0.9863–1.0779). The random-forest model had a calibration intercept of *−*0.3561 (95% CI, *−*0.4370 to *−*0.2734) and a calibration slope of 1.7750 (95% CI, 1.6928–1.8586).

**Fig. 4.**
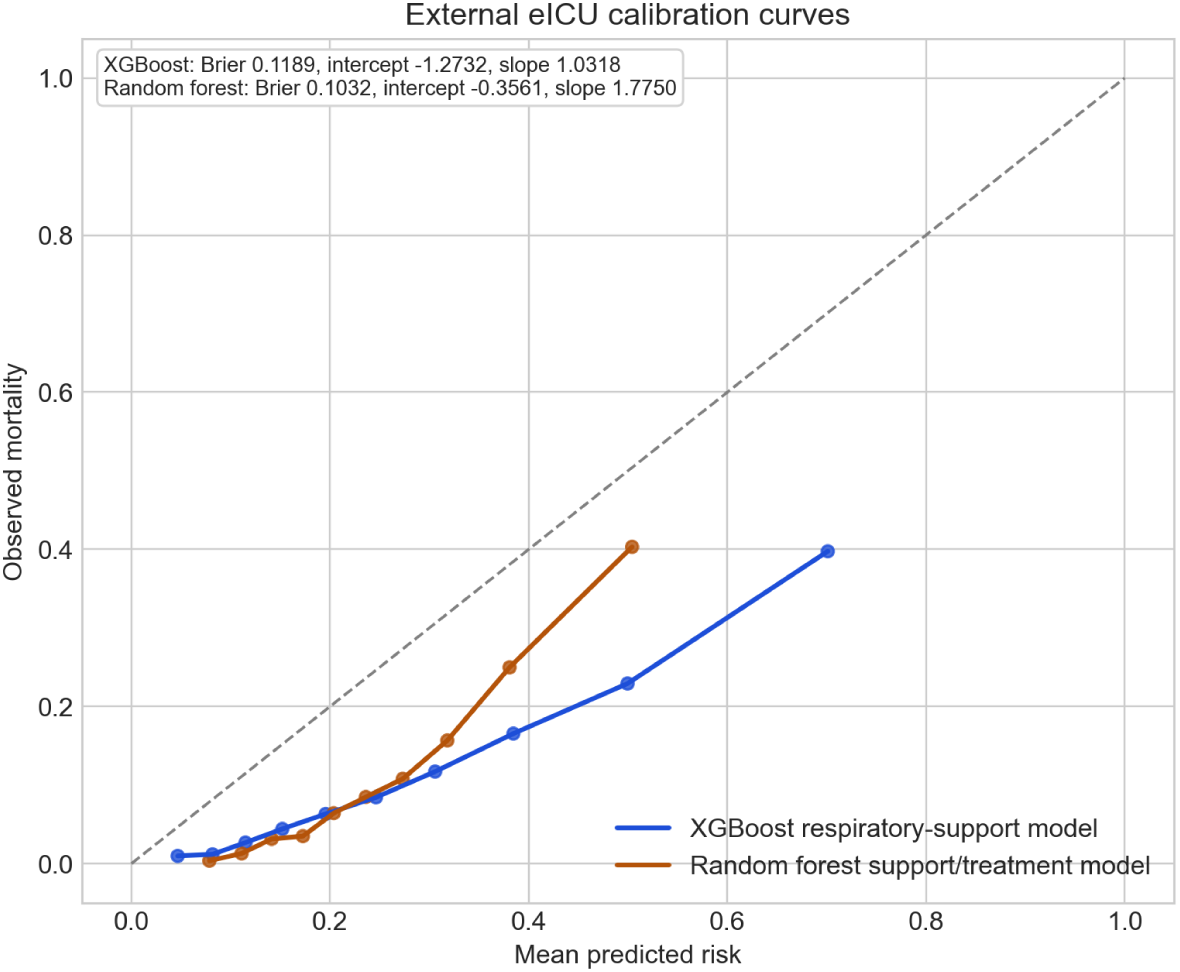
External calibration of the XGBoost and random-forest models in the eICU evaluation cohort. The diagonal reference line represents perfect agreement between predicted and observed mortality. Calibration curves were constructed using equal-frequency predicted-risk groups.

The negative calibration intercepts indicated systematic differences between predicted and observed mortality levels in eICU. The XGBoost slope was close to 1, whereas the random-forest slope was substantially greater than 1, indicating that the random-forest predictions were more compressed than the observed risk gradient. Neither model demonstrated ideal external calibration without model updating.

### 3.6 Feature importance

Native tree-based feature-importance rankings for the two model specifications are shown in Figure 5. For the XGBoost model, highly ranked predictors included tidal-volume setting count, the most recent positive end-expiratory pressure value, the most recent and mean set respiratory rates, pH, lactate, red cell distribution width, and blood urea nitrogen.

**Fig. 5.**
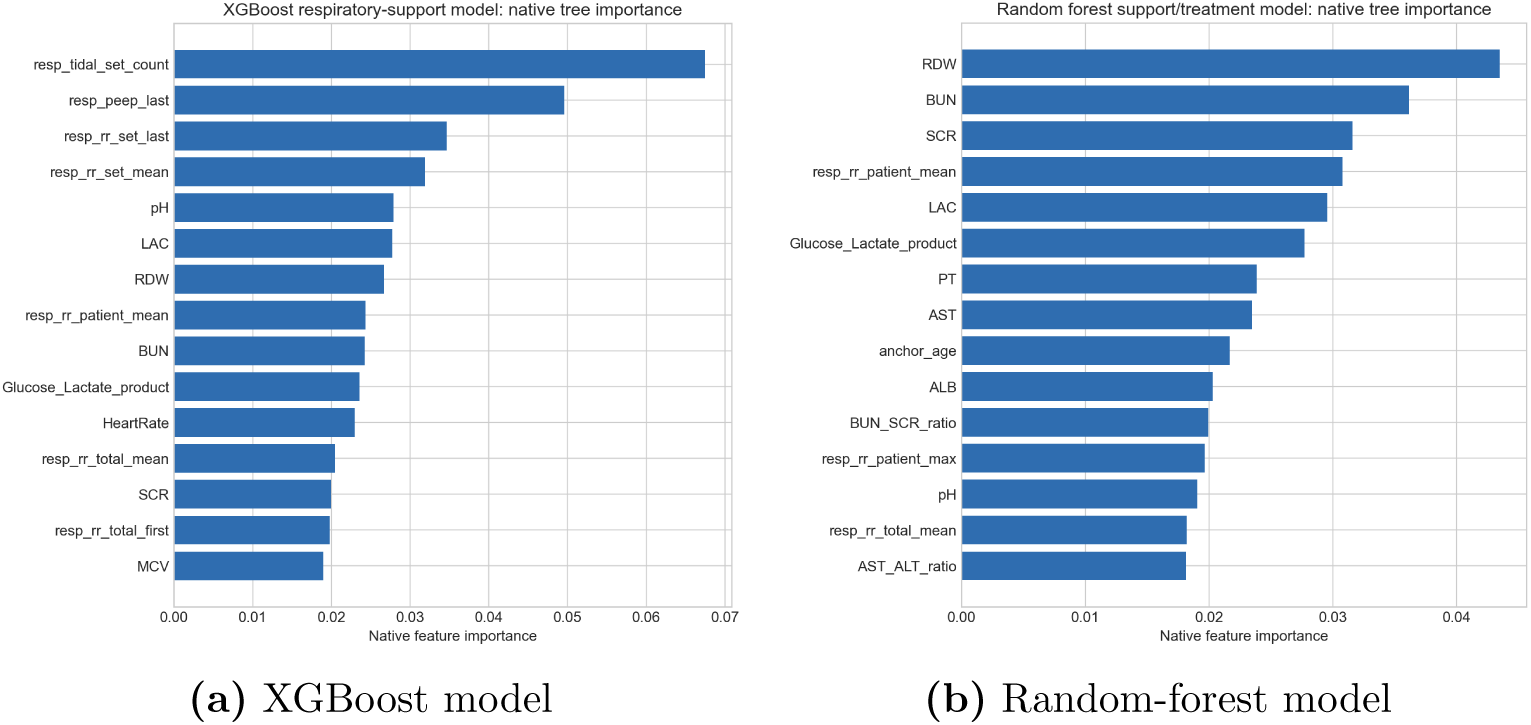
Native tree-based feature-importance rankings for **(a)** the XGBoost model using baseline and respiratory-support predictors and **(b)** the random-forest model using baseline, respiratory-support, and treatment predictors. Importance values for missingness-indicator columns were aggregated with their corresponding parent predictors. The values indicate the relative use of predictors by each fitted ensemble and do not represent signed associations or causal effects.

For the random-forest model, leading predictors included red cell distribution width, blood urea nitrogen, serum creatinine, mean recorded patient respiratory rate, lactate, the glucose–lactate product, prothrombin time, aspartate aminotransferase, and age.

Because these rankings were derived from native tree importance, they indicate the relative use of predictors within each fitted ensemble but do not provide the direction or magnitude of associations. They should not be interpreted as causal effects.

### 3.7 Decision-curve analysis

Decision-curve results for the eICU cohort are shown in Figure 6. Both models demonstrated positive net benefit relative to the treat-none strategy over portions of the low-threshold range. The random-forest model generally showed slightly higher net benefit than XGBoost across much of the low-to-moderate threshold range.

**Fig. 6.**
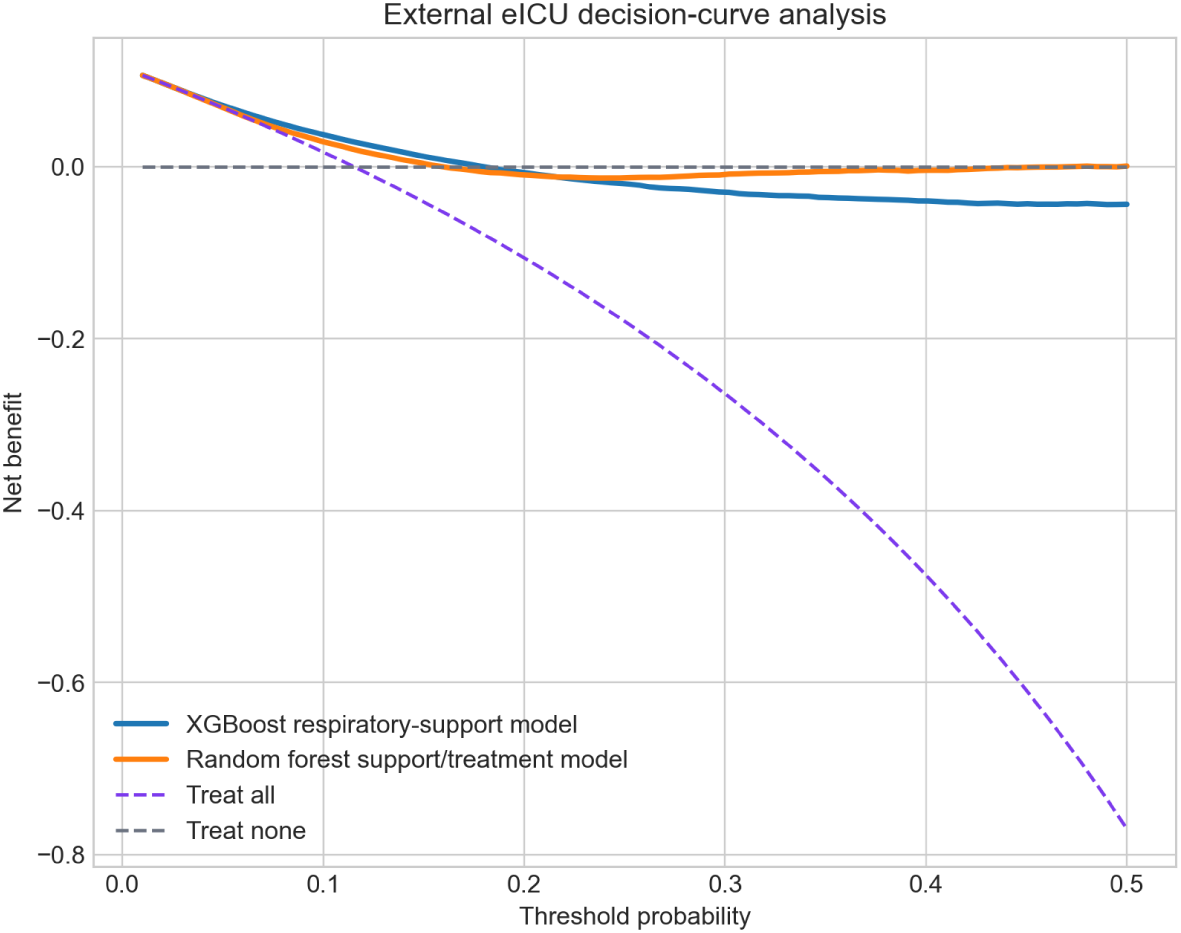
Decision-curve analysis for the XGBoost and random-forest models in the eICU evaluation cohort. Net benefit is shown across threshold probabilities from 0.01 to 0.50 together with the treat-all and treat-none reference strategies.

Net benefit decreased as the threshold probability increased, and both models approached or fell below the treat-none strategy at higher thresholds. Because no specific clinical action or decision threshold was defined in advance, these findings were interpreted as exploratory.

### 3.8 Subgroup performance

Exploratory subgroup ROC-AUC estimates for the XGBoost model are shown in Figure 7. Discrimination varied across age, sex, respiratory-support event status, and treatment-flag status. Estimates for most subgroups were broadly centered around the overall eICU ROC-AUC, although lower discrimination was observed among stays with a recorded treatment flag.

**Fig. 7.**
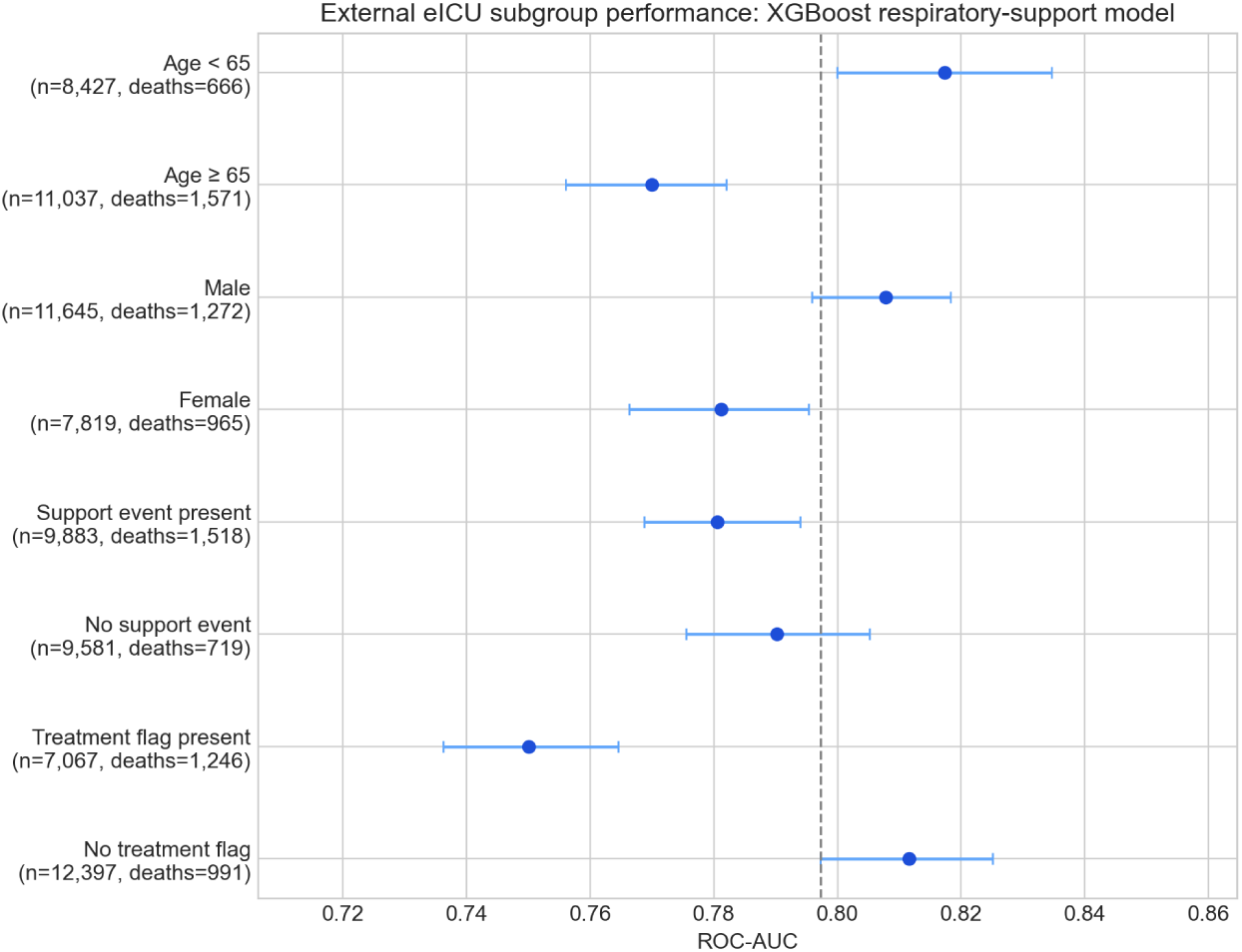
Exploratory subgroup ROC-AUC estimates for the XGBoost model in the eICU evaluation cohort. Error bars represent bootstrap 95% confidence intervals. The vertical dashed line indicates the overall eICU ROC-AUC.

These analyses were descriptive, and no formal statistical tests of interaction or differences between subgroups were performed.

### 3.9 Cross-database feature and predicted-risk distribution differences

Selected cross-database feature-distribution measures are shown in Figure 8**(a)**. Differences were observed across several respiratory-support, laboratory, and treatment-related predictors according to the population stability index, absolute standardized mean difference, and difference in missingness proportion.

**Fig. 8.**
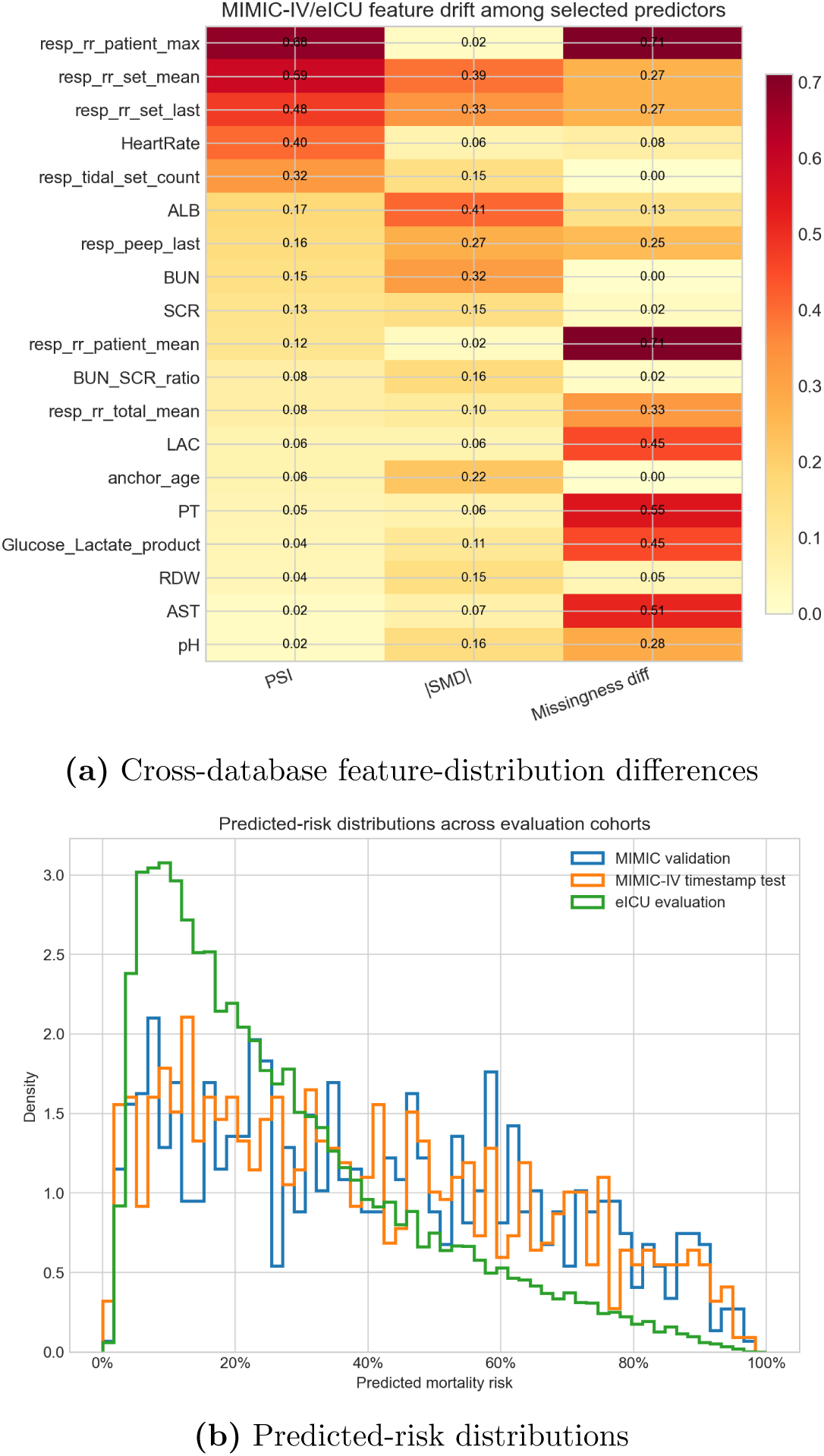
Exploratory cross-database distribution analyses. **(a)** Selected feature differences between MIMIC-IV and eICU, summarized using the population stability index, absolute standardized mean difference, and absolute difference in missingness proportion. **(b)** Predicted-risk distributions in the MIMIC-IV validation cohort, timestamp-ordered MIMIC-IV test cohort, and eICU evaluation cohort.

Predicted-risk distributions are shown in Figure 8**(b)**. Compared with the MIMIC-IV validation and timestamp-ordered test cohorts, predicted risks in the eICU cohort were more concentrated in the lower-risk range. These descriptive differences were consistent with the cross-database changes observed in mortality prevalence and external calibration.

## 4 Discussion

### 4.1 Principal findings

This study developed an ICU hour-24 landmark prediction pipeline for subsequent inhospital mortality among critically ill patients with coronary artery disease. Dynamic respiratory-support and treatment events were restricted to the first 24 hours before aggregation, models were developed in MIMIC-IV, and the final specifications were evaluated in eICU.

Beyond the point estimates, the central contribution of this study was the explicit alignment of the hour-24 landmark, predictor window, and database-specific event timestamps, together with an exploratory assessment of feature-family transport across MIMIC-IV and eICU.

Both models retained moderate-to-good discrimination in the external evaluation cohort, with ROC-AUCs of 0.7973 for XGBoost and 0.8060 for the random forest. The difference in ROC-AUC was statistically supported by paired bootstrap analysis but was small in absolute magnitude. Across repeated patient-grouped MIMIC-IV splits, the two specifications also showed similar mean validation discrimination. These findings suggest that the principal result is not the superiority of one algorithm, but the ability of timestamp-restricted first-day predictors to retain discriminatory information across the two databases.

The exploratory feature-family analyses further showed that broader feature expansion did not consistently improve cross-database performance. Respiratory-support and treatment-oriented specifications performed more consistently in eICU than feature families incorporating broader vital-sign or support/treatment extensions. However, external calibration remained imperfect, indicating that discrimination was more transportable than absolute predicted risks.

### 4.2 Comparison with prior studies

Previous disease-specific ICU mortality studies have examined dynamic or temporally summarized predictors in patients with myocardial infarction, alcoholic cirrhosis, and abdominal aortic aneurysm [14–16]. Direct numerical comparison is limited because the studies differed in patient populations, outcome definitions, prediction windows, feature construction, and validation strategies.

The present study extends this line of work by making the hour-24 information boundary an explicit part of cohort and predictor construction and by evaluating fixed model specifications across MIMIC-IV and eICU. This distinction is important because predictors derived from complete ICU or hospital stays may incorporate information recorded after the intended prediction time, producing performance estimates that do not represent the stated clinical task [9]. The landmark design used here instead defined both the eligible population and the available predictor window at ICU hour 24.

Unlike an admission-time mortality model, the resulting estimates apply only to patients who survived and remained under ICU observation through the first day. The models therefore address reassessment after 24 hours of intensive care rather than initial triage. This narrower target population should be considered when comparing performance with studies that make predictions at admission or include early deaths in the outcome population.

### 4.3 Interpretation of model and feature transport

The contribution of respiratory-support and treatment information is clinically plausible because these variables capture aspects of early respiratory failure, ventilatory requirements, treatment intensity, and physiologic instability observed during the first ICU day. Laboratory and physiologic variables such as red cell distribution width, blood urea nitrogen, creatinine, lactate, pH, and coagulation measures were also highly ranked by the fitted models. These importance rankings indicate how frequently or strongly variables were used by the tree ensembles, but they do not establish direction, causality, or clinical mechanism.

The exploratory feature-family results suggest that adding more variables may reduce rather than improve transportability when their definitions, measurement frequencies, missingness patterns, or documentation practices differ between databases. The observed feature-distribution and missingness differences between MIMIC-IV and eICU are consistent with this interpretation, although the analyses cannot determine which specific source of heterogeneity caused the performance changes.

The two final model specifications should therefore be viewed as complementary rather than as evidence that the random forest was uniformly superior. The random forest achieved a modestly higher external ROC-AUC and a lower Brier score, but its calibration slope of 1.7750 indicated compressed predictions. In contrast, the XGBoost model had a slope closer to 1 but a substantially negative calibration intercept. The difference in mortality prevalence between MIMIC-IV and eICU may have contributed to the intercept shift, but prevalence differences alone do not explain all aspects of calibration performance.

These results reinforce the need to distinguish discrimination from calibration when evaluating transported prediction models [32]. A model may preserve the ordering of higher- and lower-risk patients while failing to preserve accurate absolute risk estimates. Neither model should therefore be used with unchanged probability thresholds in another clinical setting without local recalibration and further evaluation.

### 4.4 Implications for model development and clinical use

From a model-development perspective, the findings emphasize that prediction time, timestamp eligibility, and cross-database feature definitions can be as important as algorithm choice. The repeated MIMIC-IV analyses showed that both algorithms had higher apparent training discrimination than validation discrimination, particularly the random forest. This training–validation gap suggests that model complexity and training fit should not be interpreted as evidence of generalizability.

The external evaluation provides stronger evidence regarding transport between MIMIC-IV and eICU, but it does not establish generalizability to all ICUs. In particular, database-specific documentation practices may affect whether an absent support or treatment event represents true non-exposure, incomplete recording, or differences in data organization. Predictor dictionaries and timestamp rules should therefore be reviewed and harmonized before models are transferred to a new database or health system.

The decision-curve analysis suggested positive net benefit over portions of the low-threshold range, but no specific clinical action or decision threshold was defined in advance. These findings should therefore be interpreted as exploratory rather than as evidence of clinical utility. Potential applications might involve supporting structured risk review or identifying patients for closer assessment after the first ICU day, but the present analysis does not establish a ready-to-use clinical decision rule.

### 4.5 Strengths, limitations, and future directions

This study has several strengths. It used an explicit hour-24 landmark, applied database-specific timestamp restrictions before feature aggregation, separated MIMIC-IV development from eICU evaluation, grouped repeated observations at the patient level during validation, and estimated external uncertainty using subject-clustered bootstrap resampling. Performance was evaluated using discrimination, calibration, probabilistic accuracy, decision curves, subgroup analyses, and descriptive measures of cross-database feature differences, consistent with current prediction-model reporting principles [24].

Several limitations should also be considered. First, the study was retrospective and depended on the accuracy and completeness of de-identified critical-care databases. Second, coronary artery disease and coronary heart disease were identified using diagnosis-code criteria, which may not fully correspond to coronary disease known at the intended prediction time.

Third, eICU represented only one external database. The findings therefore demonstrate transport between MIMIC-IV and eICU rather than general transportability across hospitals, countries, or electronic health-record systems. Fourth, the feature-family comparisons performed after the primary external benchmark were exploratory. They should not be interpreted as independent confirmatory validations or as definitive evidence that particular feature families will transport better in other datasets.

Fifth, the hour-24 landmark excludes patients who died or left the ICU during the first 24 hours. The findings do not apply to early mortality or admission-time triage. Sixth, several potentially relevant hemodynamic variables, including harmonized systolic, diastolic, and mean arterial pressure measurements, were not included in the final cross-database feature set. Seventh, both models showed training–validation performance gaps, particularly the random forest, and external calibration was not ideal.

Finally, native tree-based feature importance does not describe the direction of predictor effects or establish causal relationships. Future work should evaluate the pipeline in additional independent cohorts, develop a fully specified cross-database predictor dictionary, examine local recalibration, and connect decision thresholds to clearly defined clinical actions. Prospective or site-level temporal evaluation would be required before integration into clinical workflows.

## 5 Conclusions

This study developed and externally evaluated an ICU hour-24 landmark prediction pipeline for subsequent in-hospital mortality in critically ill patients with coronary artery disease. Models using timestamp-restricted first-day respiratory-support and treatment information retained ROC-AUCs of approximately 0.80 when transported from MIMIC-IV to eICU.

The findings indicate that explicit alignment of the prediction landmark, feature window, and event timestamps can support more realistic cross-database evaluation. Exploratory analyses further suggested that narrower and more consistently harmonized feature families may transport more reliably than broader feature expansions. However, external calibration remained imperfect, and the observed performance applies specifically to patients who survived and remained in the ICU through hour 24.

These models should therefore be interpreted as cross-database prediction benchmarks rather than deployment-ready probability tools. Additional external evaluation, local recalibration, and prospective assessment tied to clearly defined clinical actions would be required before clinical implementation.

## Supporting information

Supplementary File 1: Algorithms S1 and S2

## Data Availability

MIMIC-IV and the eICU Collaborative Research Database are available to credentialed researchers through PhysioNet under their respective data-use agreements. The datasets cannot be redistributed by the authors. Supplementary File 1 contains pseudocode describing the timestamp-restricted feature-construction and model-evaluation workflows. Analysis code supporting the findings of this study is available from the corresponding author on reasonable request, subject to the applicable PhysioNet data-use requirements.

https://physionet.org/content/mimiciv/

https://physionet.org/content/eicu-crd/

## Supplementary information

Supplementary File 1 contains Algorithm S1 for timestamp-restricted hour-24 feature construction and Algorithm S2 for model development, robustness assessment, and external validation.

## Abbreviations

CAD: Coronary artery disease
CHD: Coronary heart disease *CI* Confidence interval
DCA: Decision-curve analysis
eICU: eICU Collaborative Research Database
ICU: Intensive care unit
MIMIC-IV: Medical Information Mart for Intensive Care IV
PEEP: Positive end-expiratory pressure
PR-AUC: Precision–recall performance summarized using average precision
PSI: Population stability index
ROC-AUC: Area under the receiver operating characteristic curve
SD: Standard deviation
SMD: Standardized mean difference

## Declarations

### Ethics approval and consent to participate

This retrospective secondary analysis used de-identified data from MIMIC-IV and the eICU Collaborative Research Database. The collection and sharing of MIMIC-IV were reviewed by the Institutional Review Board of Beth Israel Deaconess Medical Center, which approved the data-sharing initiative and granted a waiver of informed consent. The eICU Collaborative Research Database contains de-identified health data and was accessed through PhysioNet after completion of the required credentialing, human-subjects research training, and data-use agreements. No direct patient contact occurred, and the authors did not have access to directly identifiable patient information.

### Consent for publication

Not applicable.

### Authors’ contributions

SP, YS, JS, MH, and NK contributed equally to this work. SP performed data curation, feature engineering, formal analysis, model development, validation, visualization, and preparation of the original manuscript draft. YS contributed to methodological review, validation of the analytical and reporting framework, interpretation of the results, journal-oriented restructuring, visualization, and substantial revision of the manuscript. JS, MH, and NK contributed to study development, methodological review, interpretation of the findings, and critical revision of the manuscript.

GP and KA contributed to clinical interpretation of the findings and critical revision of the manuscript. MP contributed to study conceptualization, methodology, supervision, project administration, interpretation of the results, and critical revision of the manuscript. All authors read and approved the final manuscript.

## Acknowledgements

The authors acknowledge the creators and maintainers of MIMIC-IV, the eICU Collaborative Research Database, and PhysioNet for making these research resources available.

