## Supplementary File 1: Algorithms S1 and S2 for "ICU Hour-24 Landmark Prediction of In-Hospital Mortality in Critically Ill Patients With Coronary Artery Disease: Development in MIMIC-IV and External Validation in eICU"

Supplementary Algorithms for
ICU Hour-24 Landmark Prediction of In-Hospital Mortality in Critically Ill Patients With Coronary Artery Disease: Development in MIMIC-IV and External Validation in eICU

Sakshie Pathak, Janet Sanjaya, Yong Si, Mohammadsaeed Haghi,
Nausin Kudrot, Greg Placencia, Kamiar Alaei, and Maryam Pishgar

This supplementary file provides pseudocode for the two principal components of the analysis workflow: timestamp-restricted feature construction and model development with internal robustness assessment and external validation. The pseudocode is intended to clarify the order of operations; the full methodological definitions are provided in the main manuscript.

**Algorithm S1** Timestamp-restricted hour-24 feature construction from raw event tables

**Input:** MIMIC-IV cohort table $C_M$ and eICU cohort table $C_E$
**Input:** MIMIC-IV event tables $\mathcal{D}_M$ and eICU event tables $\mathcal{D}_E$
**Input:** Prediction landmark $\tau = 24$ hours
**Output:** Harmonized feature matrices $X_M$ and $X_E$

Identify adult ICU stays with CAD or CHD according to the database-specific diagnosis-code phenotype.
Exclude stays with age $< 18$ years, ICU length of stay $< \tau$, invalid timing information, or unavailable in-hospital mortality outcome.
Restrict the landmark population to patients who are alive and remain under ICU observation at hour $\tau$.
Create one analysis row for each eligible ICU stay in each database.
Initialize binary support/treatment indicators and event-count variables to zero.
**for all** MIMIC-IV event tables $D \in \mathcal{D}_M$ **do**
    Join events to the corresponding ICU admission time.
    Retain only events satisfying
        $\text{ICU intime} \leq \text{event time} \leq \text{ICU intime} + 24 \text{ h}.$
    Map source-specific event names to prespecified harmonized feature groups.
    Aggregate retained events by ICU stay using applicable summaries: any event, count, first, last, minimum, maximum, mean, and change.
**end for**
**for all** eICU event tables $D \in \mathcal{D}_E$ **do**
    Retain only events satisfying
        $0 \leq \text{event offset}_{\text{min}} \leq 1440.$
    Map source-specific event names to the corresponding harmonized feature groups.
    Aggregate retained events by ICU stay using the same applicable summary rules.
**end for**
Left-join all aggregated feature tables to the full eligible cohort in each database.
Encode the absence of a recorded support or treatment event as zero for binary indicators and event counts.
Preserve unavailable numeric summaries as missing values for subsequent fitting-data-based imputation.
Align MIMIC-IV and eICU to the shared predictor definitions required by each model specification.
**return** $X_M$ and $X_E$.

**Algorithm S2** Model development, robustness assessment, and external validation

**Input:** Harmonized MIMIC-IV feature matrix $X_M$ and outcome $Y_M$

**Input:** Harmonized eICU feature matrix $X_E$ and outcome $Y_E$

**Output:** Internal and external evaluation results for the two final model specifications

1: Create a subject-grouped MIMIC-IV development partition:
   (i) reserve 20% of subjects as the held-out internal test set;
   (ii) split the remaining 80% of subjects 75:25 into training and validation subsets;
   (iii) retain all eligible ICU stays from each subject within the assigned subset.

2: Define each subject's first eligible ICU admission time as
   $\texttt{first\_intime} = \min(\texttt{intime})$ within `subject_id`.

3: Sort subjects by `first_intime` and then by `subject_id`.

4: Assign the earliest 70% of subjects to the timestamp-ordered training set and the latest 30% to the timestamp-ordered test set.

5: Estimate imputation parameters using the corresponding MIMIC-IV fitting data only.

6: Apply the fitted preprocessing parameters unchanged to the associated validation and test subsets.

7: Train candidate XGBoost and random-forest models using MIMIC-IV data only.

8: Select the XGBoost baseline-plus-respiratory-support specification according to the MIMIC-IV validation ROC-AUC criterion.

9: Retain the random-forest baseline-plus-respiratory-support-plus-treatment specification as the secondary MIMIC-IV-defined model.

10: Keep all eICU rows, outcomes, predictions, and performance estimates unavailable during model fitting, hyperparameter selection, and initial feature-family selection.

11: **for** $r = 1$ to 30 **do**

12: Repeat the subject-grouped MIMIC-IV partitioning procedure using the prespecified split proportions and a different random seed.

13: Re-estimate preprocessing parameters from the repeat-specific training subset.

14: Refit both final model specifications on the repeat-specific training subset.

15: Evaluate each model on the repeat-specific validation subset.

16: Record training ROC-AUC, validation ROC-AUC, validation average precision, and validation Brier score.

17: **end for**

18: Summarize the repeated-split performance distributions.

19: Re-estimate preprocessing parameters using all 4,341 eligible MIMIC-IV analysis stays.

20: Refit both final model specifications on the full MIMIC-IV analysis cohort.

21: Apply the fitted preprocessing pipelines and models unchanged to the eICU external-validation cohort.

22: Estimate external uncertainty using 1,000 subject-clustered bootstrap resamples of eICU.

23: Within paired bootstrap replicates, compare random forest minus XGBoost for ROC-AUC and Brier score.

24: Report ROC-AUC, average precision, Brier score, calibration intercept, calibration slope, calibration curves, decision curves, feature importance, and subgroup performance.

25: After the primary eICU benchmark, conduct exploratory feature-family transport and cross-database distribution analyses.
